# Clinical implementation of routine whole-genome sequencing for hospital infection control of multi-drug resistant pathogens

**DOI:** 10.1101/2022.05.02.22273921

**Authors:** Brian M Forde, Haakon Bergh, Thom Cuddihy, Krispin Hajkowicz, Trish Hurst, E. Geoffrey Playford, Belinda C. Henderson, Naomi Runnegar, Julia Clark, Amy V Jennison, Susan Moss, Anna Hume, Hugo Leroux, Scott A Beatson, David L Paterson, Patrick NA Harris

## Abstract

**Background:** Prospective whole-genome sequencing (WGS)-based surveillance may be the optimal approach to rapidly identify transmission of multi-drug resistant (MDR) bacteria in the healthcare setting.

**Materials/methods:** We prospectively collected methicillin-resistant *Staphylococcus aureus* (MRSA), vancomycin-resistant enterococci (VRE), carbapenem-resistant *Acinetobacter baumannii* (CRAB), extended-spectrum beta-lactamase (ESBL-E) and carbapenemase-producing Enterobacterales (CPE) isolated from blood cultures, sterile sites or screening specimens across three large tertiary referral hospitals (2 adult, 1 paediatric) in Brisbane, Australia. WGS was used to determine *in silico* multi-locus sequence typing (MSLT) and resistance gene profiling via a bespoke genomic analysis pipeline. Putative transmission events were identified by comparison of core genome single nucleotide polymorphisms (SNPs). Relevant clinical meta-data were combined with genomic analyses via customised automation, collated into hospital-specific reports regularly distributed to infection control teams.

**Results:** Over four years (April 2017 to July 2021) 2,660 isolates were sequenced. This included MDR gram-negative bacilli (n=293 CPE, n=1309 ESBL), MRSA (n=620) and VRE (n=433). A total of 379 clinical reports were issued. Core genome SNP data identified that 33% of isolates formed 76 distinct clusters. Of the 76 clusters, 43 were contained to the three target hospitals, suggesting ongoing transmission within the clinical environment. The remaining 33 clusters represented possible inter-hospital transmission events or strains circulating in the community. In one hospital, proven negligible transmission of non-multi-resistant MRSA enabled changes to infection control policy.

**Conclusions:** Implementation of routine WGS for MDR pathogens in clinical laboratories is feasible and can enable targeted infection prevention and control interventions.

**Summary:** We initiated a program of routine sequencing of multi-drug resistant organisms. A custom analysis pipeline was used to automate reporting by incorporating clinical meta-data with genomics to define clusters and support infection control interventions.

## INTRODUCTION

Healthcare-associated infections (HAIs) are common and associated with significant morbidity and excess healthcare-related cost [1-3]. Each year in Australia, more than 165,000 patients experience HAIs [4]. Increasing rates of antimicrobial resistance (AMR) exacerbates the impact of these infections [5] and adds considerable cost burden to the healthcare system [3]. In Australia, 30-day mortality rates for hospital-onset vancomycin resistant *Enterococcus* (VRE) and methicillin-resistant *Staphylococcus aureus* (MRSA) bloodstream infections were 20% and 14.9% respectively [6], with mortality from extended-spectrum beta-lactamase (ESBL)-producing *E. coli* bloodstream infections up to 18.6% [7]. Traditional molecular methods used to describe potential genetic relationships between organisms lack resolution, may be time consuming and are not readily available outside specialist laboratories [8]. Whole genome sequencing (WGS) is now established as the optimal method to analyse outbreaks and explore transmission dynamics of bacterial pathogens [9] and could conceivably act as a frontline tool in the analysis and management of most pathogens that represent a threat to human health [10].

There has been limited diagnostic laboratory capacity to track pathogens causing these infections or detect transmission events in the healthcare setting. This study aimed to develop a clinical WGS workflow, able to prospectively detect transmission events before they become established, guide infection prevention and control interventions and respond to outbreaks.

## MATERIALS AND METHODS

### Setting

The project was initiated in three tertiary-referral hospitals within metropolitan Brisbane, Australia, to provide regular WGS reports to infection control teams. Sequencing services were also available on request for regional facilities served by the state-wide laboratory network (Pathology Queensland). Recommendations for the management of multi-resistant organisms (MROs) in Queensland Health facilities are based on state-wide guidelines [11].

#### Sample collection

Patient inclusion criteria were defined as:

1. Any patient hospitalised at three tertiary care facilities (Royal Brisbane and Women’s Hospital [RBWH], Princess Alexandra Hospital [PAH] and Queensland Children’s Hospital [QCH]) with a MRO screening culture positive for any of the following: MRSA, VRE, ESBL-producing Enterobacterales (ESBL-E), carbapenem-resistant *Acinetobacter baumannii* (CRAB) or carbapenemase-producing Enterobacterales (CPE).
2. Any patient admitted at RBWH, PAH or QCH with a positive culture for a target pathogen (ESBL/CPE, MRSA, VRE or CRAB), from Sterile Site [Blood, tissues, fluids or CSF] cultures.
3. Any patient hospitalised at outlying metropolitan Brisbane hospitals or regional Queensland Health Facilities with CRAB or CPE referred to the central referral laboratory for molecular testing. WGS was available on request for other bacterial species or resistance phenotypes in the context of suspected outbreaks or unexplained resistance phenotypes (e.g. suspected carbapenemase production, without carbapenemase genes detected by PCR).

### Data collection

Basic patient demographic data (age, hospital / ward location, dates of admission) and sample data (date of collection, species identification, resistance profile) were automatically entered into a REDCap electronic data capture system [20] using HL7 V2 messaging originating from the Pathology Queensland laboratory information system (AUSLAB, Citadel Health, Melbourne). The data were then integrated into the genomic analysis pipeline (Figure 1).

**Figure 1:**
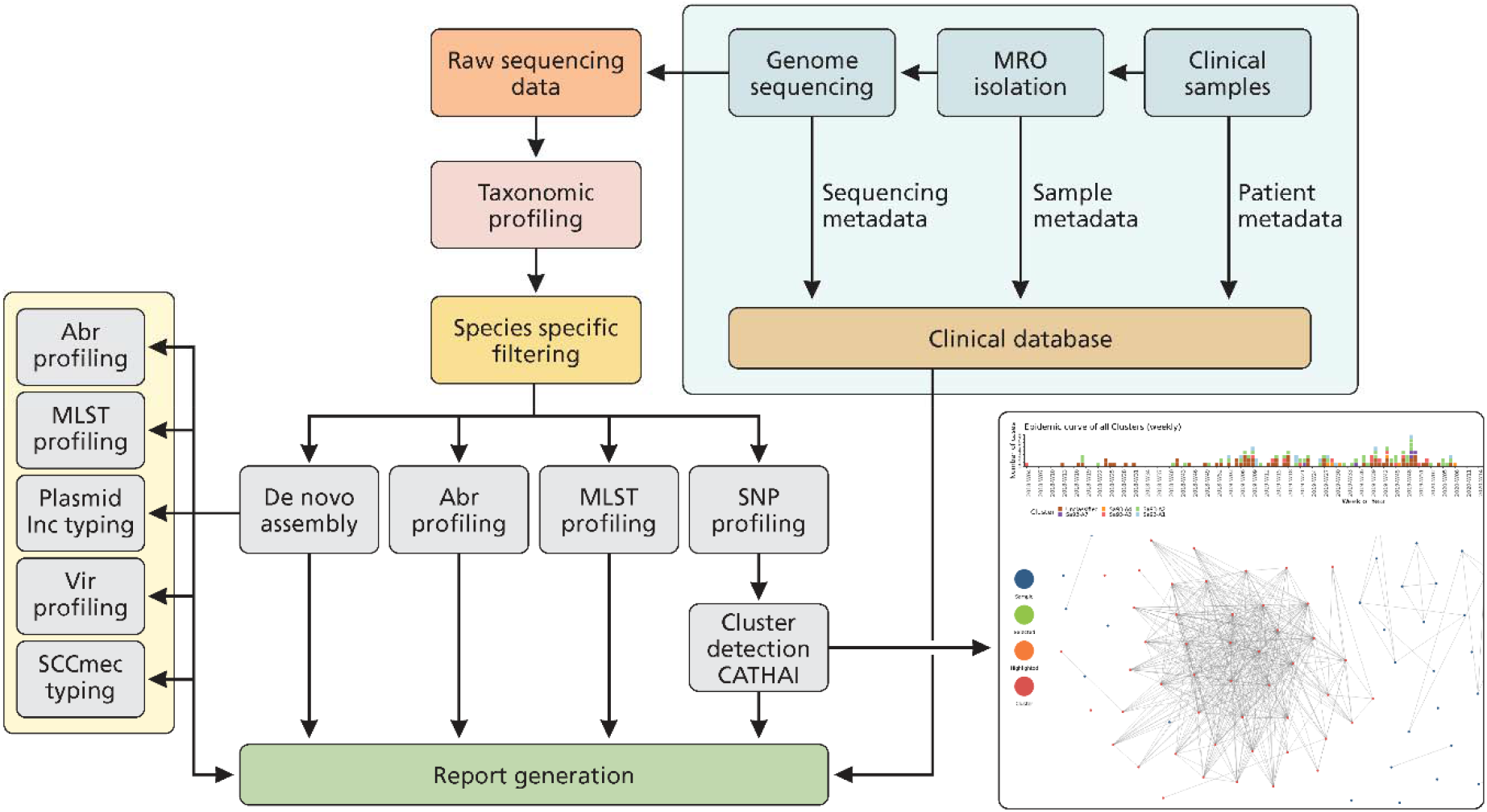
Schematic representation of the workflow, including sample processing, clinical meta-data collection, SnapperRocks analysis pipeline, cluster visualisation and clinical report generation. MRO = multi-resistant organism; Abr = Antibiotic resistance; MLST = multi-locus sequence typing; SNP = single nucleotide polymorphism; Vir = virulence; SCC*mec* = staphylococcal chromosomal cassette *mec*; CATHAI = Cluster Analysis Tool for Healthcare-Associated Infections.

### Microbiology

All collected isolates we subjected to culturing, identification, and reporting according to routine laboratory protocols at Pathology Queensland (Supplementary material). Acceptable screening samples for MRSA included skin/mucosal swabs, respiratory samples or urine; for VRE, swabs (rectal/perineal) or faeces; for ESBL/CPE/CRAB, skin/mucosal swabs, urine or faeces.

### Whole genome sequencing

All patient isolates were submitted for WGS (Illumina Nextseq 500; 150bp paired-end) at the Queensland Health Forensic Scientific Services (FSS) laboratory in weekly batches (Supplementary material).

### Genome analysis pipeline

Genomic analysis was undertaken using a custom, in-house microbial genomic analysis pipeline, SnapperRocks (https://github.com/FordeGenomics/SnapperRocks). SnapperRocks uses Conda (https://anaconda.org/) for tool management, and the NextFlow workflow engine (https://www.nextflow.io/) to achieve reproducibility and easy deployment across personal computers or high-performance computing clusters (Figure 1). Pipeline workflow is discussed in detail in the supplementary material.

### SNP profiling and Clustering

Sequence Reads for each isolate were mapped to a strain-specific representative reference genome using SNPdragon (*https://github.com/FordeGenomics/SNPdragon*). In brief, trimmed reads were aligned to a reference using BWA-mem. Samtools [12] mpileup was used to calculate per base coverage and freebayes (https://github.com/freebayes/freebayes) to call variants.

Pair-wise core genome SNP differences were used to identify clusters of genetically similar isolates where local transmission was likely. Three or more Isolates were deemed to form a probable transmission cluster if the pair-wise SNP differences between them was no more than 5 core genome SNPs/Mb. Isolates were defined as community-associated if they were collected within 2 days of hospital admission and hospital-associated if collected >2 days post-admission [13]. The mean collection time of all isolates within a cluster was used to determine if a particular cluster was community associated (CA) or hospital associated (HA).

## RESULTS

### Sample collection

Between 19th April 2017 and 1st July 2021, a total of 2,660 bacterial isolates were included, cultured from samples obtained from the participating hospitals. The isolates were collected from 2336 patients, with 259 patients providing multiple isolates. A further 64 isolates were obtained from environmental sources within participating hospitals. Of the 2596 patient-derived isolates, 1535 (58.8%) were collected from screening specimens (e.g. rectal swabs), 278 (10.6%) from invasive clinical infections (blood cultures [n=277] and cerebrospinal fluid [n=1]), 772 (29.6%) from non-invasive clinical infections (Table S1). Samples were collected weekly, with an average of 8 samples (min=0; max=39) sent for whole genome sequencing (WGS) each week.

### *In silico* taxonomic profiling and multi-locus sequence typing (MLST)

MRO isolates collected over the course of this project included representatives of 50 bacterial species from 22 genera. Dominant species accounted for 93% of collected isolates (Table 1) [14]. The remaining 173 isolates we assigned to 37 different species, including *Citrobacter freundii* (n=14), *Serratia marcescens* (n=8) and *Pseudomonas aeruginosa* (n=30) (Table S1).

**Table 1:**
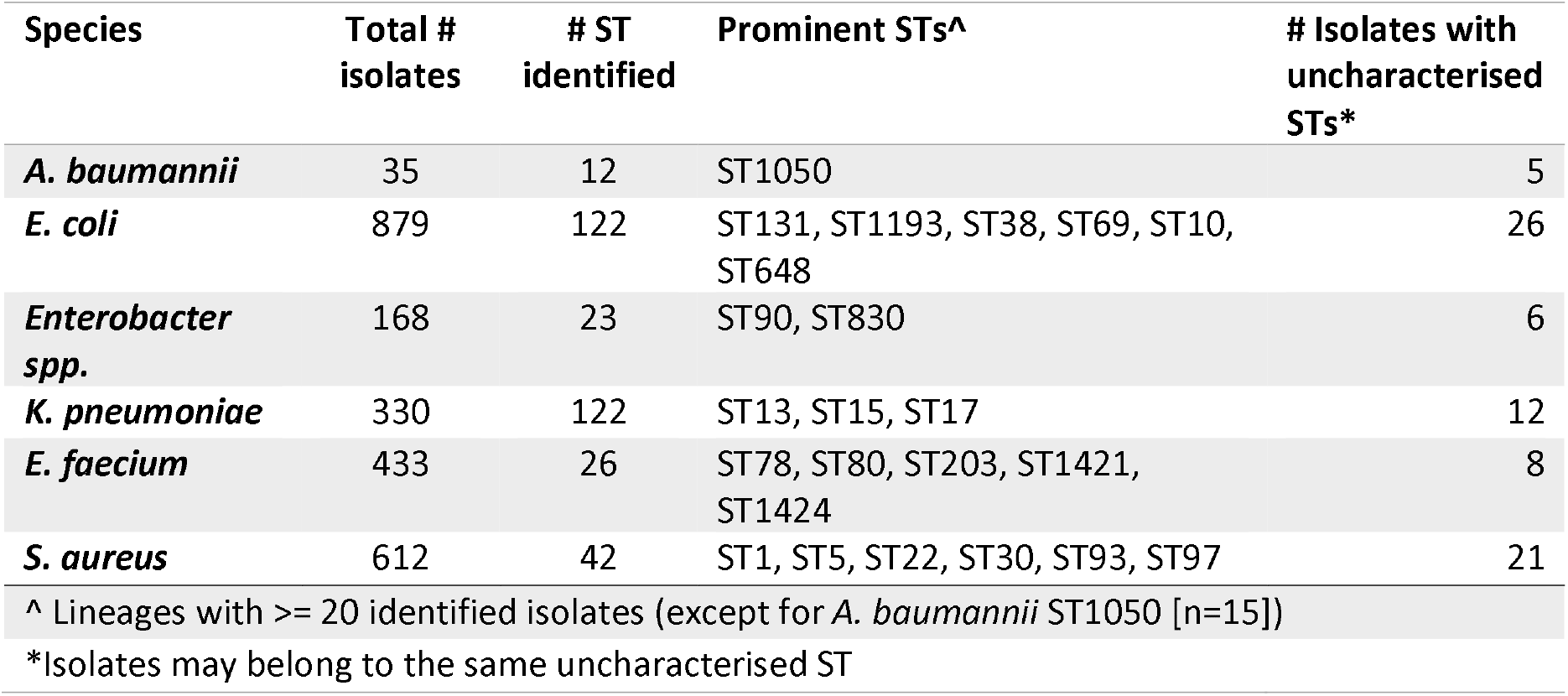
Lineage diversity among sequenced isolates

| Species | Total # isolates | # ST identified | Prominent STs<sup>^</sup> | # Isolates with uncharacterised STs* |
| --- | --- | --- | --- | --- |
| *A. baumannii* | 35 | 12 | ST1050 | 5 |
| *E. coli* | 879 | 122 | ST131, ST1193, ST38, ST69, ST10, ST648 | 26 |
| *Enterobacter spp.* | 168 | 23 | ST90, ST830 | 6 |
| *K. pneumoniae* | 330 | 122 | ST13, ST15, ST17 | 12 |
| *E. faecium* | 433 | 26 | ST78, ST80, ST203, ST1421, ST1424 | 8 |
| *S. aureus* | 612 | 42 | ST1, ST5, ST22, ST30, ST93, ST97 | 21 |
| <sup>^</sup> Lineages with >= 20 identified isolates (except for *A. baumannii* ST1050 [n=15]) |  |  |  |  |
| * Isolates may belong to the same uncharacterised ST |  |  |  |  |

MLST profiling revealed high levels of species diversity among 5 most abundant species (Table 1). *E. coli* were dominated by ST131 (n=276; 37.3%), which accounted for 10% of all clinical isolates (Figure 2). In contrast, there was no single dominant lineage among *K. pneumoniae*. However, lineages associated with the spread of carbapenem resistance were present; specifically, ST11, ST15, ST101 and ST258 [15]. Dominant *Enterobacter* spp. lineages (*E. hormaechei* ST90 and ST830) and *A. baumannii lineages* (ST1050) were related to previously described outbreaks [16]. *S. aureus* were mainly ST93, a community associated clone frequently identified in Queensland [17, 18]. All prominent *E. faecium* lineages belonged to clonal complex 17 (CC17), responsible for a significant burden of healthcare-associated infections [19].

**Figure 2:**
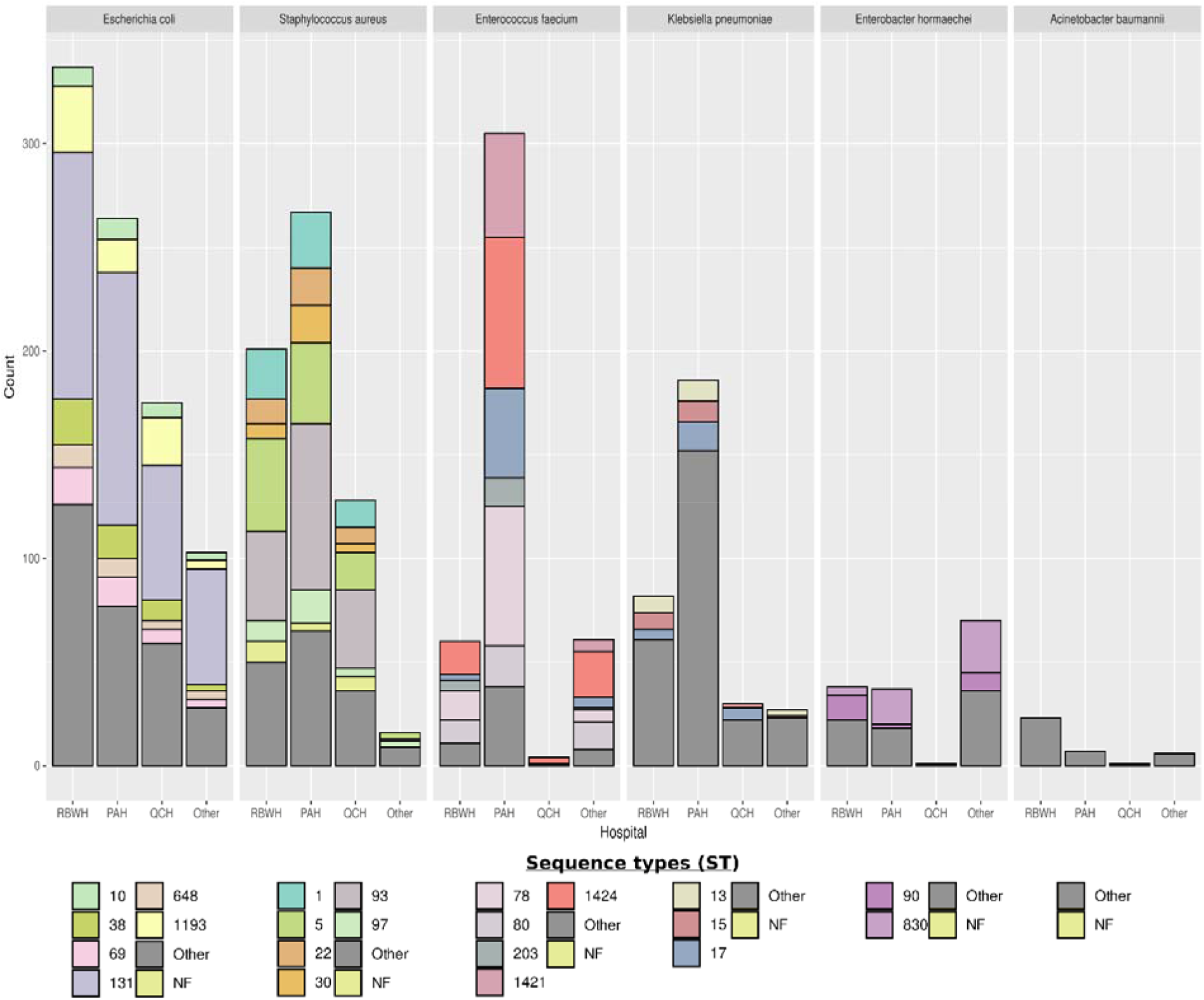
Distribution of clonal lineages among the 5 most prevalent species. Lineages with <20 representative members are classified as “other” and isolates whose sequence type had not previously been defined at the time of writing were designated as “NF”.

### Antibiotic resistance genes

In addition to ESBL and other beta-lactamases, GNBs carried a diverse range of genes conveying resistance to numerous classes of antibiotics, including aminoglycosides, tetracyclines, sulphonamides and quinolones (Table S2).

Carriage of carbapenamase genes was identified in 289 isolates from 21 Enterobacterales species (Figure 3; Table S1). *bla*_IMP-4_ was most commonly observed among CPEs (n=170), consistent with previous reports [20, 21]. Five different NDM homologues were identified: *bla*_NDM-1_ (n=9), *bla*_NDM-4_ (n=2), *bla*_NDM-5_ (n= 25), *bla*_NDM7_ (n=1) and *bla*_NDM-9_ (n=2) (Table S1 & Table S2). Other carbapenemases included OXA-types (e.g. OXA-48 [n=10], OXA-23 [n=31], OXA-181 [n=11]), KPC-2 (n=5) and IMI-1 (n=1). Isolates carrying both NDM and OXA carbapenemases were detected in 7 patients (Table S1). Two carbapenemase-producing *Pseudomonas aeruginosa* carrying IMP-1 and DIM-1 were identified. Other carbapenemases intrinsic to less common species were occasionally identified (e.g. GOB in *Elizabethkingia anophelis*).

**Figure 3:**
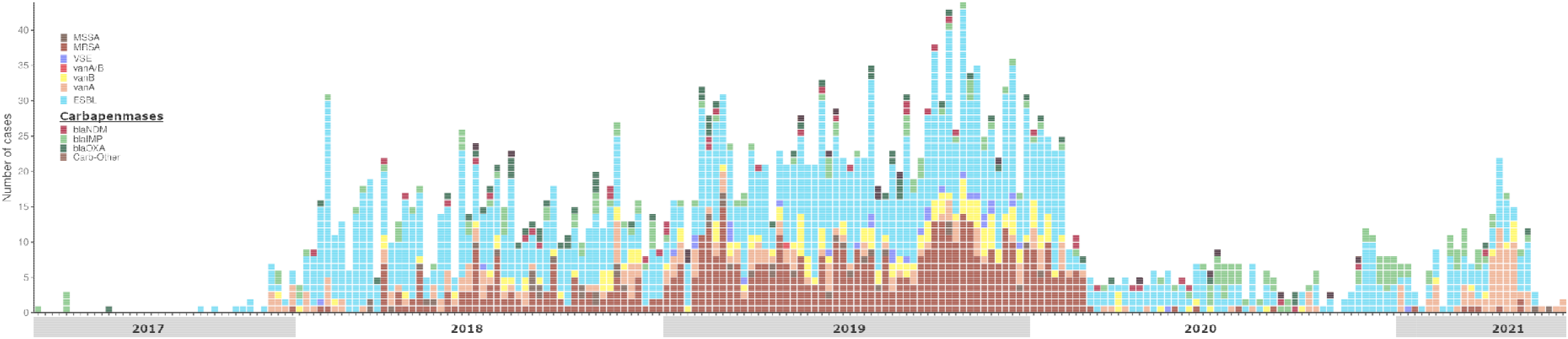
Temporal distribution and frequency of resistance-associated genotypes identified among clinical Isolates. Isolates are grouped by week of collection from April 2017 to July 2021.

The *mec* gene (*mecA*), which mediates methicillin resistance, was identified in 92.5% (569/615) of *S. aureus* isolates (Figure 3). The absence of *mec* (n=46 isolates) usually reflected errors in phenotypic testing due to calculated microbroth MICs and fluctuations around interpretative breakpoints.

In enterococci, *vanA* (n = 239) was more common than *vanB* (n=159), reflecting both global and national trends [22]. However, rates may reflect sampling bias towards the more resistant *vanA* gene. Additionally, 33 vancomycin-susceptible *E. faecium* (VSEfm) were sequenced to determine whether these were clonally related to circulating healthcare-acquired VRE (Figure 3).

### Clustering and Transmission

Characterising the genomic relationship (SNP distance) between isolates revealed that, 33% (864/2660) of samples were found to cluster with two or more isolates of the same clonal lineage (Figure 4). Overall, a total of 76 clusters were identified across the six most frequently identified species (Table 2).

**Table 2:** Clustering of predominant species

| Species | Clustered isolates | #Clusters | STs with clusters | Maximum Cluster Size (Cluster ID) |
| --- | --- | --- | --- | --- |
| *A. baumannii* | 18 | 3 | ST1050, ST451, ST229 | 14 isolates (Ab1050-A1) |
| *E. coli* | 249 | 27 | ST10, ST38, ST69, ST73, ST88, ST127, ST131, ST141, ST185, ST349, ST410, ST457, ST1193, ST4553 | 76 isolates (Ec131-A8) |
| *Enterobacter spp.*<sup>^</sup> | 60 | 4 | ST90, ST513, ST599, ST830 | 23 Isolates (Eh415-B1) |
| *K. pneumoniae* | 67* | 7 | ST13, ST22, ST133, ST348, ST429, ST711, ST1412 | 17 isolates (Kp13-A1) |
| *E. faecium* | 289 | 19 | ST17, ST78, ST80, ST203, ST555, ST780, ST1421, ST1424 | 74 isolates (Ef1421-A2) |
| *S. aureus* | 149 | 17 | ST5, ST30, ST72, ST78, ST88, ST93, ST97, ST239 | 51 isolates (Sa93-A2) |
| * Includes 19 *K. michiganensis* isolates from a neonatal SCU outbreak (KmNA-A1). <sup>^</sup> Includes: *E. bugandensis*, *E. cloacae*, *E. hormaechei* |  |  |  |  |

**Figure 4:**
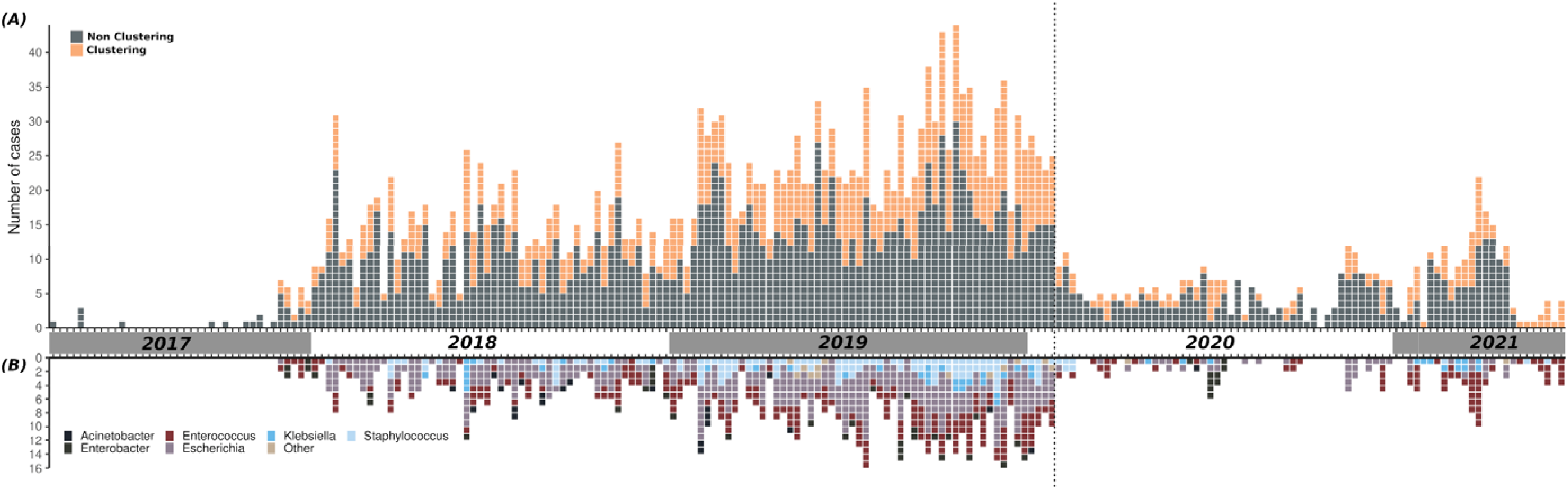
**(A) Epidemiological curve of all isolates collected during this study.** Isolates are grouped by week of collection from April 2017 to July 2021. Dotted line indicates a change in sample collection with a focus on a targeted strategy, prioritising organisms of greatest concern (e.g. CPE, CRAB, VRE, ESBL-*K. pneumoniae*) or invasive clinical isolates of other pathogens (e.g. ESBL-*E. coli*, MRSA). **(B) Epidemiological curve of clustering isolates coloured by genus.** Only the top 5 genera are shown, with all other clustering isolates classified as “Other”.

Average cluster size was 4.5 isolates, with 77.6% of clusters containing <10 isolates. Two clusters, Ab1050-A1 and Eh90-A2, were linked to previously described nosocomial outbreaks of *A. baumannii* [16] and *E. hormaechei* [20], respectively. Finally, 79 single transmission events (2 isolate clusters) were also detected, without further onward transmission (Table S1).

The majority (43/76) of our defined clusters were geographically confined to a single hospital. The remaining 31 clusters had a wider geographic distribution with isolates collected from two or more different facilities, suggesting the possibility of inter-facility transmission networks.

As a maker of healthcare-associated (HA) transmission the time between hospital admission and sampling was inspected. For 35 of our clusters, the mean period between patient admission and isolate collection (DeltaAS) was found to be ≤2 days, suggesting that these clusters could have arisen through community rather than nosocomial transmission (Figures S1-S6, Supplementary material), although we cannot rule the possibility of transfer from other unsampled healthcare facilities. Collectively, *Escherichia* and *Staphylococcus* species, accounted for 70% of suspected CA-clustering with lower rates of suspected community clustering among other frequently identified species (Figure 5).

**Figure 5:**
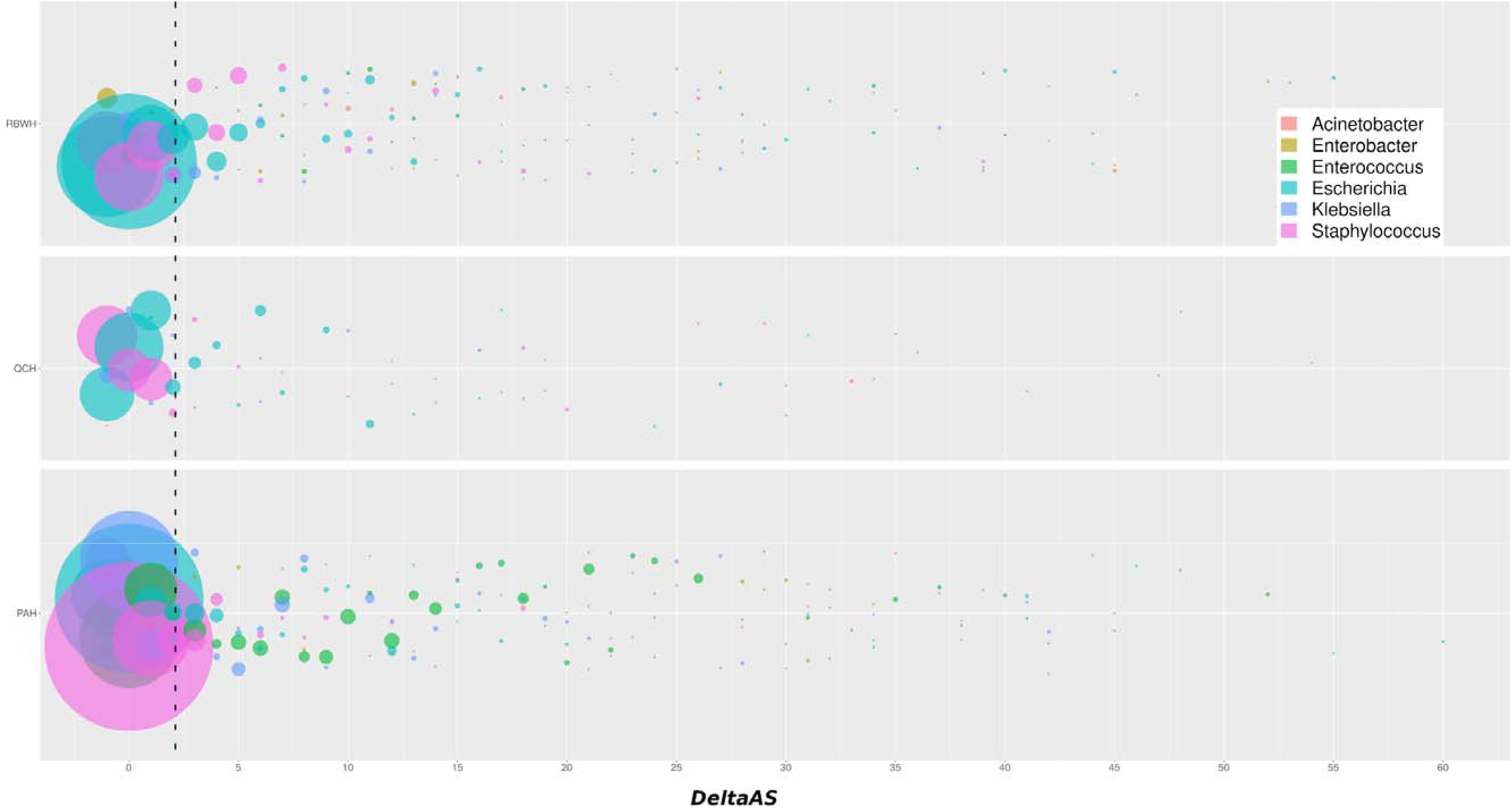
Community versus healthcare-association by genus. Clustering isolates are coloured by genus, as per the legend, and scaled by number of isolates. Dashed line indicates a sample collection time of 2 days post admission (DeltaAS=2). RBWH=Royal Brisbane & Women’s Hospital; QCH=Queensland Children’s Hospital; PAH=Princess Alexandra Hospital.

### Genomics reporting and integration with infection control

During the study period, a total of 379 genomics reports were issued to infection control teams. The median time for sample processing (collection to referral for WGS) was 10 days. 41% of isolates arrived for sequencing ≥11 days post collection and included >300 “historical” isolates collected >30 days (min 30; max 1025) before processing (Figure S7B, Supplementary material). The median turnaround time for sequencing was 7 days, with 29% of samples taking ≥8 days (min 8; max 48) (Figure S7C, Supplementary material). The median turnaround time for genomic analysis and report generation was 6 days (min 0; max 120), with 43% of samples taking ≥8 days (Figure S7D, Supplementary material). Overall, the median duration from sample collection to final genomic reporting was 33 days (+/-1 standard deviation: 21 to 45 days), with the fastest reports delivered within 10 days (Figure 6; Figure S7A, Supplementary material).

**Figure 6:**
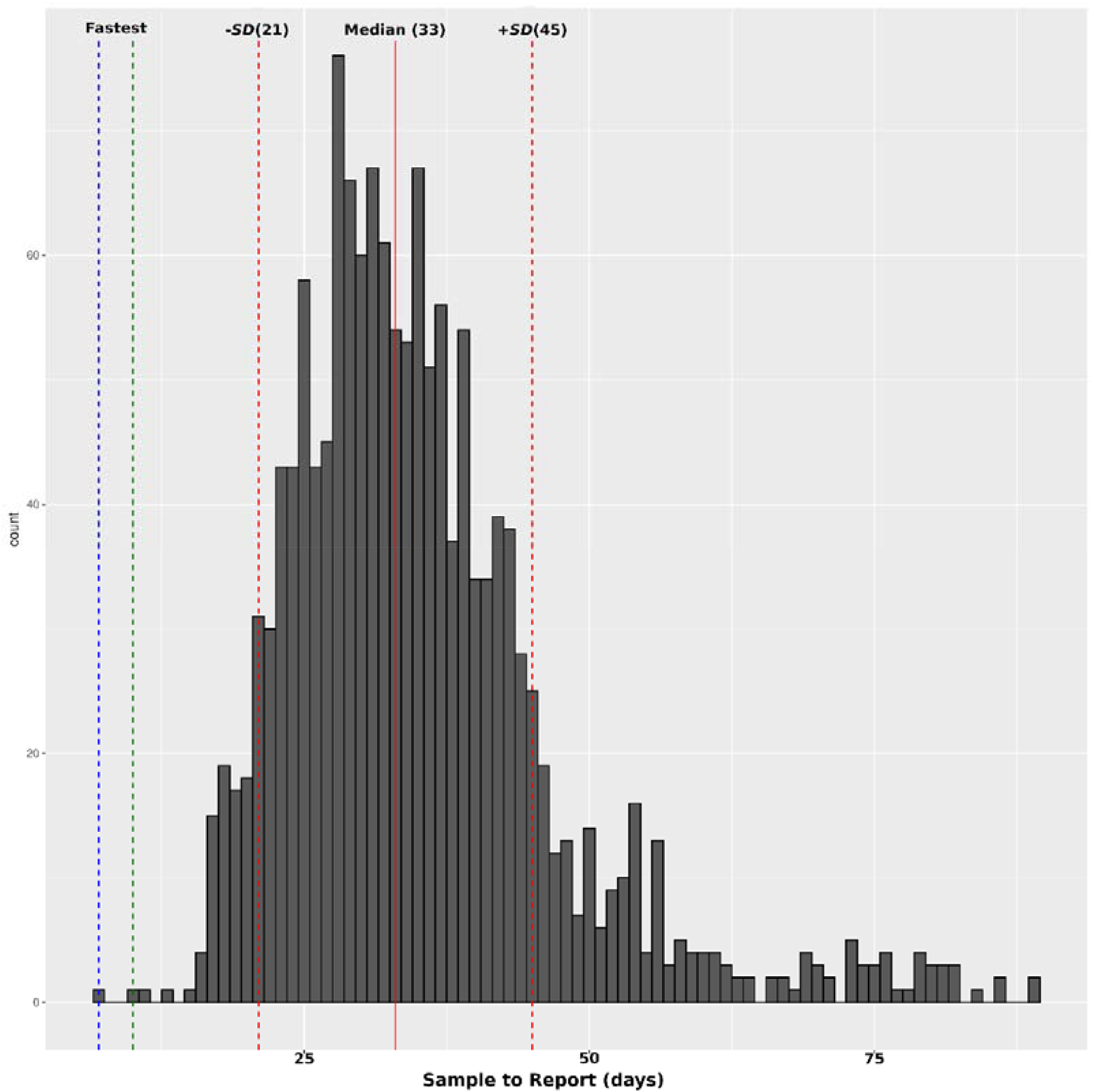
**Turn-around times for genomic reports** from day of sample collection to report generation with a cut off of 90 days. Green and blue dashed lines represented the achieved and possible reporting periods, 10 and 7 days respectively.

Reports were discussed during regular hospital infection control committee meetings and presented to hospital administrators routinely and in response to outbreaks. The design of the reports (see supplementary material) aimed to concisely summarise the key characteristics of collected organisms, alert the team to any likely clustering (between current and/or previously sequenced isolates and established clusters) and describe key resistance genes. Additional dynamic exploration of the relationships between isolates could be achieved by accessing the online data visualisation portal CATHAI (https://cathai.fordelab.com/), which can also generate line-listings of patient meta-data and epidemic curves of defined clusters.

### Implementation of genomic surveillance into clinical practice

To enhance the translational impact of the surveillance program, we involved infection prevention and control nurse specialists with an interest in the application of genomics. Their roles were essential for linking the genomic reports to the clinical environment and ensuring suspected clusters were investigated and subsequent action occurred. Overall, the genomic surveillance program provided several key advantages when complementing routine infection control practice. Specifically, the reports allowed unequivocal determination of whether isolates were linked, and the likelihood of transmission between patients or healthcare environments. Conversely, it was useful in disproving transmission events in suspected outbreaks when isolates with similar phenotypes were identified in patients within shared locations. Critically, it allowed identification of early clusters to initiate more-targeted infection control measures (e.g. intensive screening of cluster-specific contacts, environmental sampling and cleaning of locations linked to clusters). WGS reporting also identified organisms associated with hospital outbreaks in other Australian states (e.g. ST1424 VRE) and could identify parallel clusters amongst phenotypically similar organisms, allowing more refined prioritisation of infection control resources. As Pathology Queensland provides a state-wide microbiology service, a genomics program such as this offered a networked surveillance system able to detect inter-hospital transmission events that would be otherwise invisible. Such a co-ordinated response to AMR threats has been identified as a critical step for public-health prevention [23].

## DISCUSSION

The application of genomics to characterise transmission routes and enhance infection prevention and control practices during clinical outbreaks is now well established [14, 20, 24]. However, these studies are largely retrospective, often temporally and spatially confined, and generally limited in scope with regards to species of interest. Additionally, the use of genomics as real time surveillance and infection prevention tool is rare. Here we describe the clinical implementation of continuous, genomic surveillance to identify, track and interrupt the transmission of multidrug resistant pathogenic bacteria.

Coupling genomic and epidemiological data revealed that 37 identified clusters had arisen through suspected community rather than hospital transmission events. However, comparison of SNP distances suggests that, even among CA-clusters, within-hospital transmission cannot be completely ruled out. Genomic-based transmission and clustering information supported existing practice to not use isolation procedures for patients colonised with ESBL-*E. coli*, and contributed to the decision at one hospital to remove isolation precautions for non-multi-resistant MRSA, resulting in estimated savings of $690,864 (AUD) over one year [25].

High levels of community transmission of MDR organisms is of great concern. Legislation to designate high-risk MDR organisms as notifiable to public health could bring additional resources to bear and help reduce the impact of community reservoirs of AMR. However, without a better understanding of the epidemiological factors that promote dissemination of MROs in the wider community, it is unclear what interventions would be most effective to interrupt these transmission networks.

Timely provision of reports is crucial for an effective surveillance program. While we were able to provide reports within 10 days of sample collection, the mean report turnaround time of 33 days stretches the boundaries of clinical relevance. However, delays associated with sample transport to the central laboratory, culture-based processing, the lack of onsite or dedicated WGS infrastructure, continuous development of the analysis pipeline, transfer of isolates to an off-site sequencing facility, batched sequencing, genomic analysis and report generation all contributed to lengthy reporting periods. However, with workflow refinements and structural re-organisation many of these delays could be minimised.

Prior to commencement of our genomic surveillance program, major gram-negative outbreaks were recorded in Brisbane hospitals in 2015 (IMP-4 carbapenemase-producing *E. hormaechei* ST90), 2016 (carbapenem-resistant *A. baumannii* ST1050) and 2017 (OXA-181 carbapenemase-producing *E. coli* ST38 and ESBL-producing *K. michiganensis*) [14, 16, 20, 26]. Targeted WGS played a critical role in the resolution of these outbreaks but was largely instigated only once extensive transmission had been detected. The added health and economic benefit of WGS-enhanced interventions is supported by recent simulation modelling, which estimates a 55.9% reduction in infections and a corresponding 59.9% decrease in associated healthcare costs over the course of an outbreak [26]. Here, the identification of putative transmission clusters Ab1050-A1 and Eh90-A2, which were linked back to two of these previous outbreaks, demonstrates that continued surveillance is required to ensure that outbreaks are completely resolved. Consequently, the true benefit of WGS can only be realised when it is deployed as a prospective surveillance tool. A recent economic evaluation of our surveillance program estimated that prospective WGS will result in ∼36,000 fewer HAIs, ∼650 fewer deaths and overall cost saving of $30million/year [27].

Despite the success of this project there were limitations. Although rare, available metadata were not always consistent between sites resulting in information gaps which impacted downstream interpretations. The importance of integrating genomic and epidemiological data has been well established as the latter provides critical contextual information, including temporal and spatial dynamics. To facilitate the coupling of data in real-time, our pipeline will need to be fully integrated with Queensland Health’s laboratory information system. This in turn allows for the publication of clinically meaningful reports in actionable timeframes while ensuring that patient privacy is protected. Secondly, this prospective surveillance program focused solely on multidrug resistant bacteria. Consequently, the health and economic impacts associated with the transmission of antibiotic susceptible disease-causing organisms was not considered

## CONCLUSIONS

The routine application of prospective genomic surveillance for MROs encountered in the hospital environment is feasible and can be implemented within a clinical diagnostic laboratory. Such an approach provides early notification of isolate clustering and likely in-hospital transmission events, thus facilitating targeted infection control responses. Despite the complex challenges of WGS pipeline development, integrating appropriate computational infrastructure within existing systems in the healthcare setting and succinct communication of complex genomics information to clinicians, WGS-informed infection prevention and control strategies are likely become the new benchmark. While the costs of running such a service are considerable, they are justified by the potential savings to the health system as a whole and enhanced prevention of healthcare-acquired infections in vulnerable patients.

## Supporting information

Supplementary material - Surveillance report template

Supplementary material - Patient report template

## Data Availability

All data produced in the present study are available upon reasonable request to the authors

## Ethics

Ethical oversight was provided by The Forensic and Scientific Services Human Ethics Committee (reference HEC17_17) as a Low and Negligible Risk approval, with provision for a waiver of individual patient consent.

## Acknowledgments

We thank the scientists at Pathology Queensland and Forensic and Scientific Services for their assistance in the laboratory work and the infection control teams at the participating hospitals for their involvement and clinical input.

