## Supplementary material for "Clinical implementation of routine whole-genome sequencing for hospital infection control of multi-drug resistant pathogens"

**SUPPLEMENTARY METHODS**

**Microbiology**

MRSA screening samples were set up on CHROMID® MRSA SMART agar (bioMérieux, France), incubated at 35°C in air, and read at 24 and 48 hours. ESBL, CPE or CRAB samples were set up on CHROMID® ESBL agar (bioMérieux, France) and, for CPE requests CHROMID® CARBA SMART agar, which were then examined at 24 hrs. Combination disk testing with cefotaxime and ceftazidime +/- clavulanic acid was performed on any *E. coli* or *K. pneumoniae* that flagged as possible ESBL in the Vitek 2 Automated Expert System (AES); a ≥5 mm increase in inhibition zone with clavulanic acid phenotypically defined the presence of an ESBL [1]. If suspected CPE (based on a minimum inhibitory concentration [MIC] to meropenem > 0.25mg/L or growth on CARBA SMART agar) a test for carbapenemase production was performed (β-CARBA test; Bio-Rad, France). Isolates with growth on either side of CARBA SMART agar and/or a positive β-CARBA test were referred for an in-house multiplex real-time PCR to detect five common Australian CPE genes (NDM, IMP-4 like, VIM, KPC and OXA-48-like genes) [2]. For VRE, samples were incubated in an in-house VRE broth for 24 hours and then sub-cultured onto CHROMID® VRE agar (bioMérieux, France), and incubated at 35°C in O_2_, examined at 24 and 48 hours. *Enterococcus* isolates with a vancomycin MIC >4 mg/L were then tested by an in-house PCR for the presence of vanA/B genes. All isolates were identified to species level by MALDI-TOF (Vitek MS, bioMérieux, France) and routine antimicrobial susceptibility testing was undertaken using Vitek 2 automated broth microdilution (bioMérieux, France). Additional MICs for meropenem (in suspected CPE organisms) or glycopeptides (vancomycin/teicoplanin in suspected VRE) were performed using E-test (bioMérieux, France).

**Whole genome sequencing**

Genomic DNA (gDNA) was extracted from fresh colony growth using DSP QIAamp DNA mini kits (Qiagen, Australia) on the QiaSymphony SP and quantified by fluorometry (Quant-iT dsDNA Assay High Sensitivity Kit; Life Technologies). Extracted DNA of a concentration greater than 1ng/µL was acceptable for further processing. Paired-end DNA libraries were prepared using Nextera XT library prep kits (Illumina; Australia) and WGS was preformed using the Illumina NextSeq 500 (150 bp paired-end). Quality Control analysis involving fastQC (v0.11.5) and MultiQC (v1.1) was performed after the removal of adapter sequences by Trimmomatic V0.36 software.

**Genomic analysis**

First, the raw Illumina sequence read data for each isolate was quality trimmed using trimmomatic (version 0.36)[3] removing low quality bases and Illumina adapter sequences. Contaminated samples were identified, and taxonomic labels assigned using Kraken 2 (version 2.0.7-beta)[4]. In brief, quality trimmed sequence read data for each isolate against were screened against a RefSeq database[5] comprised exclusively of bacterial genomes. The initial bacterial genome database was created on 19/10/2017 and updated in 2018 (26/11/2018) and 2019 (11/10/2019), respectively. Taxonomic classifications were used to assign references for *in silico* MLST and SNP profiling.

Next quality trimmed sequence reads for each isolate were assembled using Spades (version 3.14.1)[6] with default parameters. Following assembly low quality contigs (contigs length < 100bp; coverage < 20X) were removed.

Multi-locus sequence typing (MLST) of isolate raw reads and assembled quality filtered contigs was performed using srst2 (version 0.2.0)[7] and mlst (version 2.16.4) (<https://github.com/tseemann/mlst>), respectively, and typing schemes available on PubMLST (<https://pubmlst.org> ). For each isolate typing schemes were chosen based on their assigned taxonomic label. *In silico* resistance gene profiles were determined for each isolate by screening the sequence reads and draft assembled genomes for each isolate against the NCBI resistance gene database using srst2 and abricate (version 0.9.8) (<https://github.com/tseemann/abricate>), respectively. Only genes with a minimum of 70% nucleotide sequence identity and 90% sequence coverage were reported. *SCCmec*-type was determined using a modified version of SCCmecFinder (<https://cge.cbs.dtu.dk/services/SCCmecFinder/>) updated to use BLAST+ plus several bug fixes and a curated database of *SCCmec* locus nucleotide sequences. Our modified version of SCCmecFinder is available from github (https://github.com/FordeGenomics).

**SNP profiling and Clustering**

Reference genomes were chosen based on isolate taxonomic labels assigned using kraken 2. Post-filtering of variant calls was performed to report high-confidence SNP positions passing the following filters: Alternative allele fraction >= 0.75, Per base depth >= 10, Mean mapping quality >= 30, Read balance >= 0.05.

Additionally, SNPs occurring in cliff regions and clusters were removed. A cliff is a region of rapidly changing read depth and is calculated by applying a linear trend line over a 10bp sliding window. Regions where the slope is <= 3 or >= 3 and the fit of the line (R2) is >= 0.7 are labelled as cliffs and masked. Clusters are defined as >= 3 SNPs in a 10bp window. A presence/absence matrix of the high-confidence SNPs positions was then populated using all SNPs for each sample.

**SUPPLEMENTARY FIGURES**

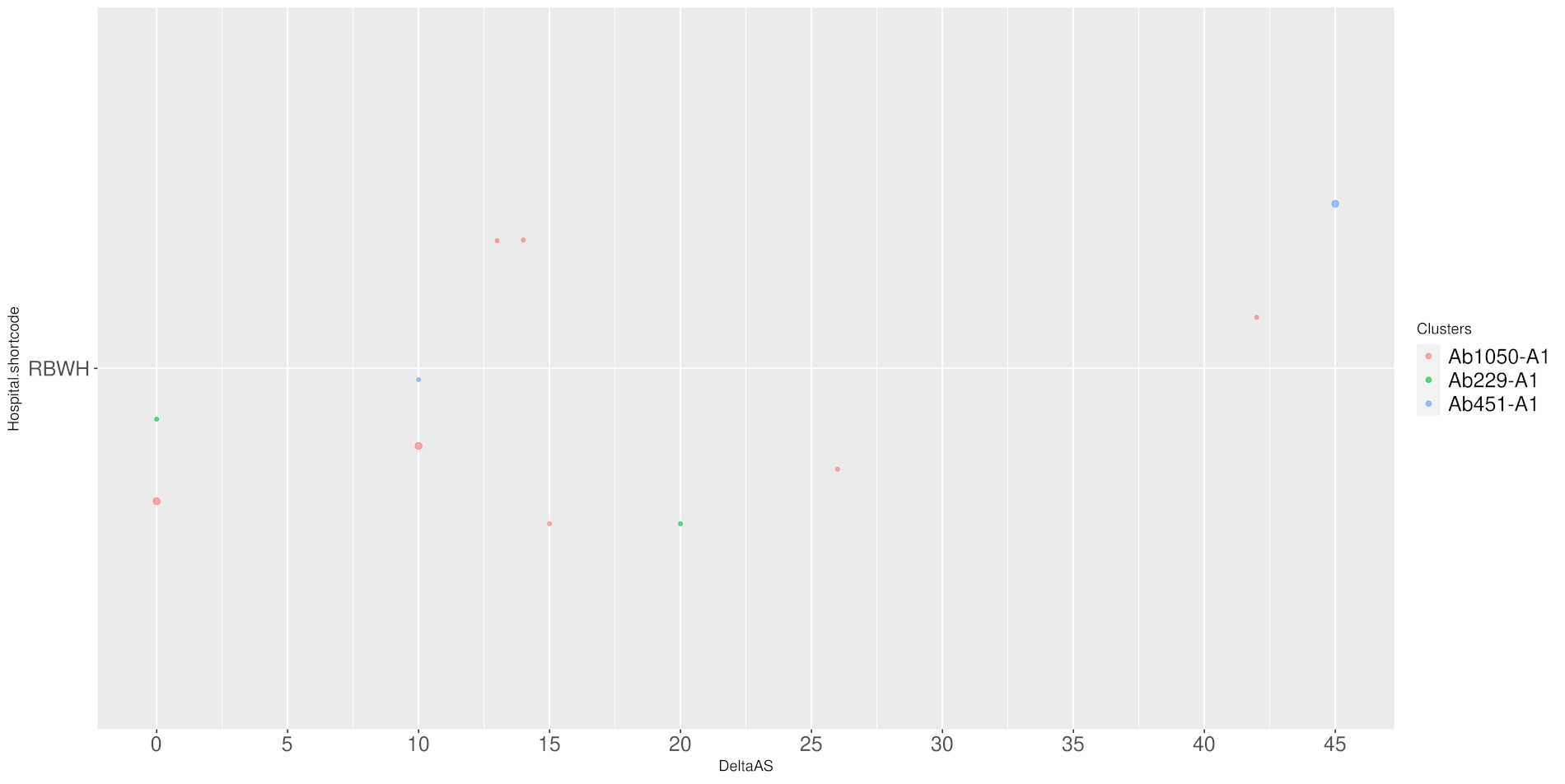

Figure S1: Patient sampling, days post admission for A. baumannii

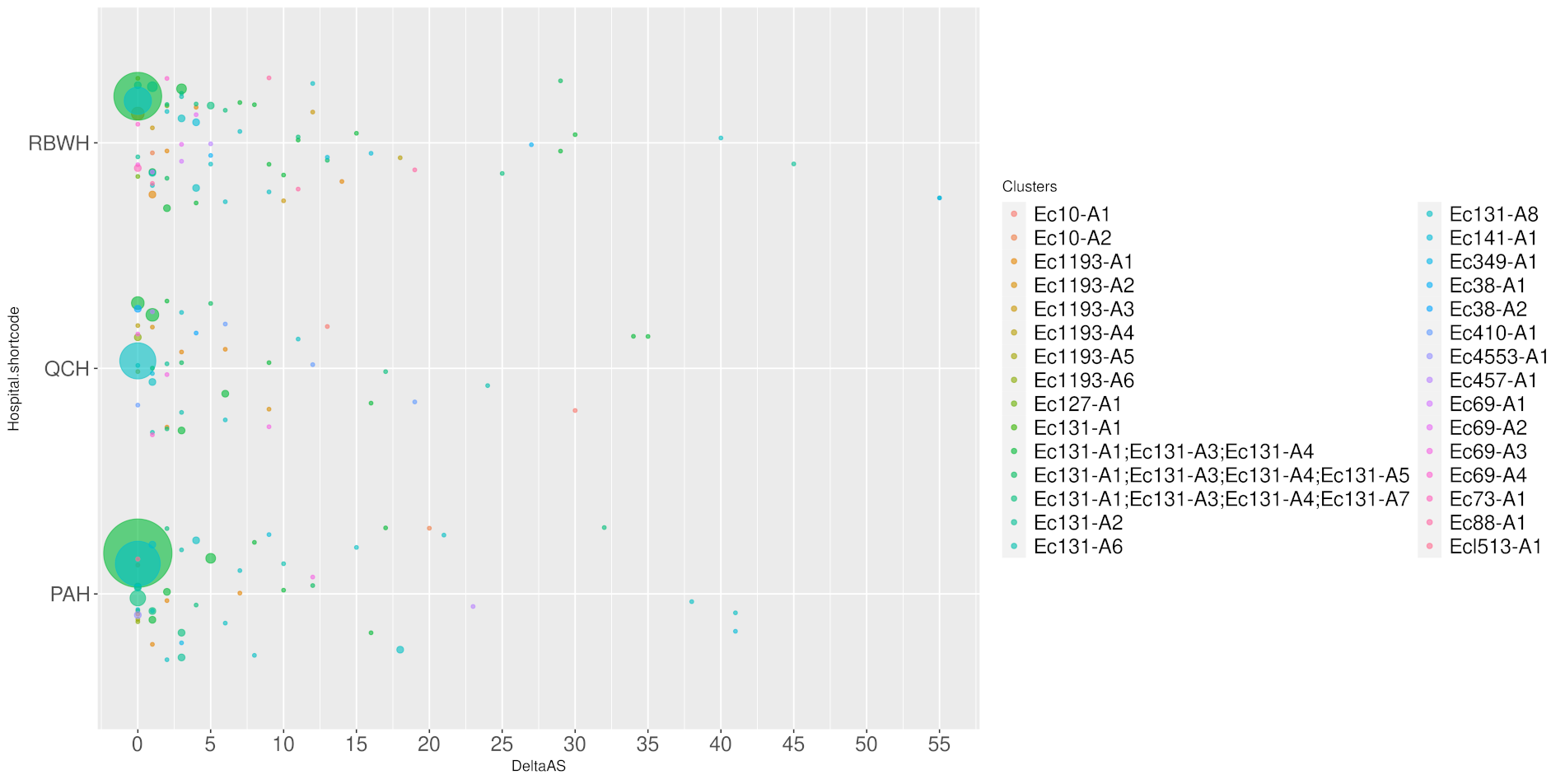

Figure S2: Patient sampling, days post admission- E. coli.

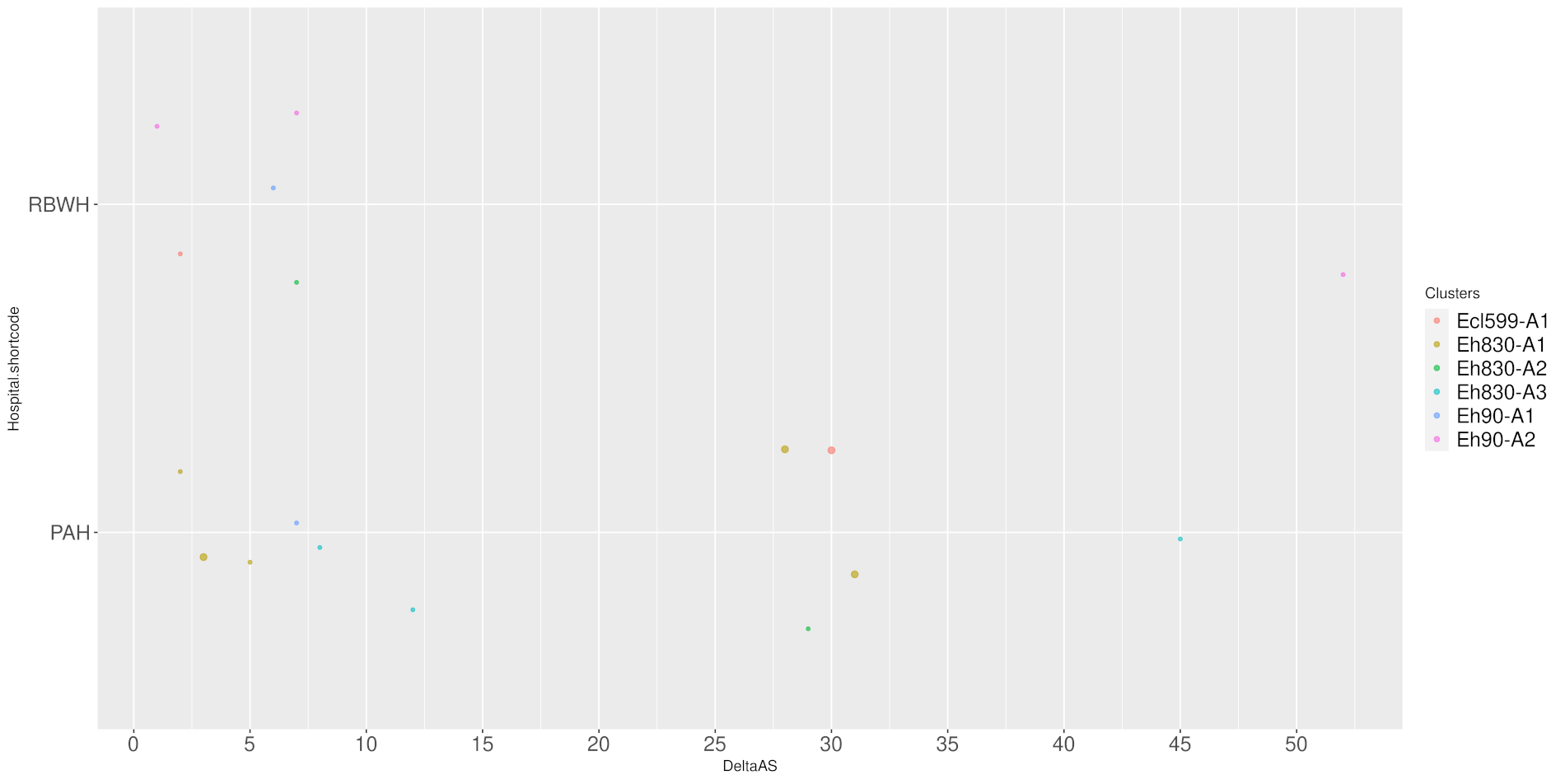

Figure S3: Patient sampling, days post admission - Enterobacter species.

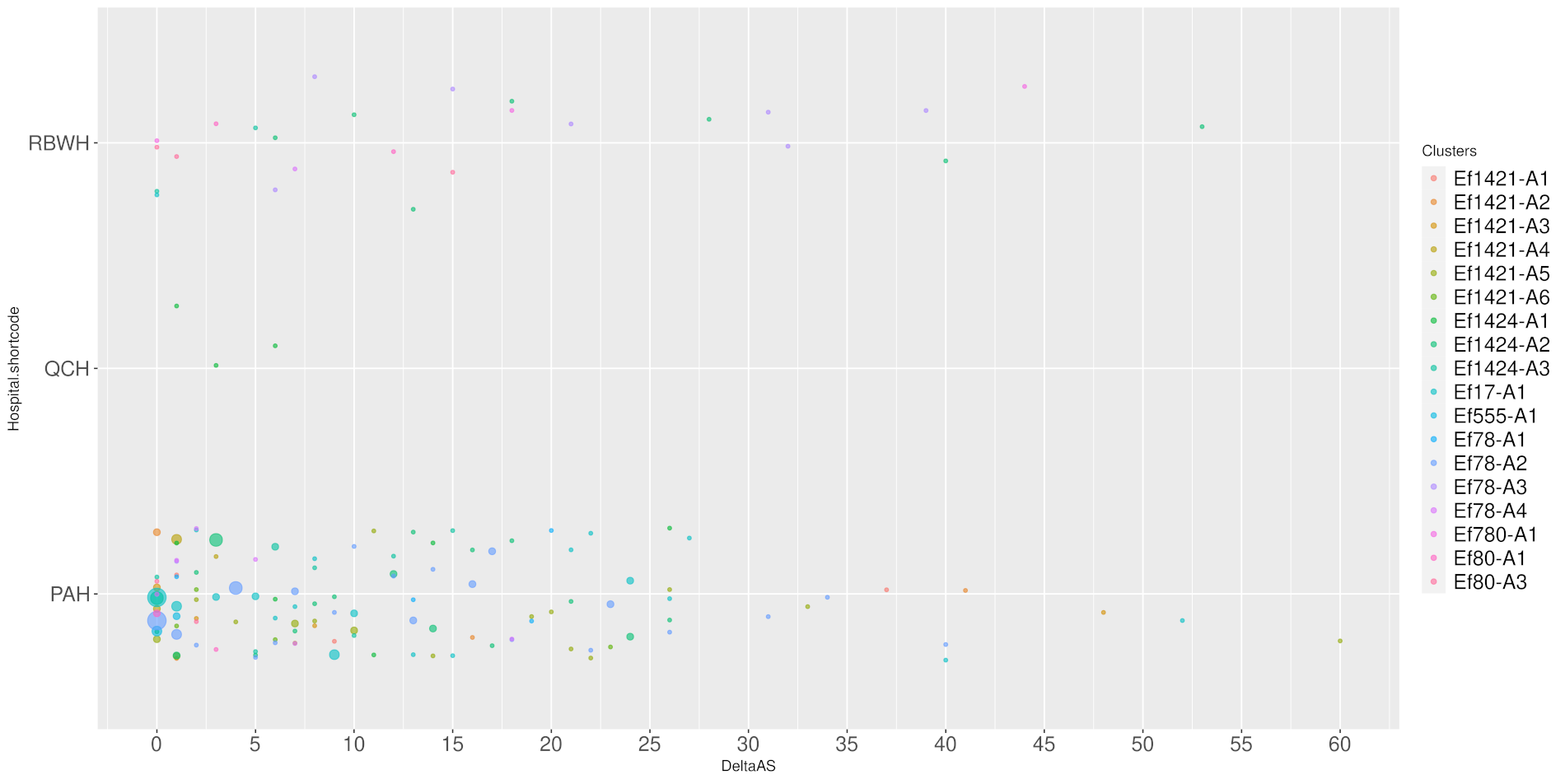

Figure S4: Patient sampling, days post admission - E. faecium.

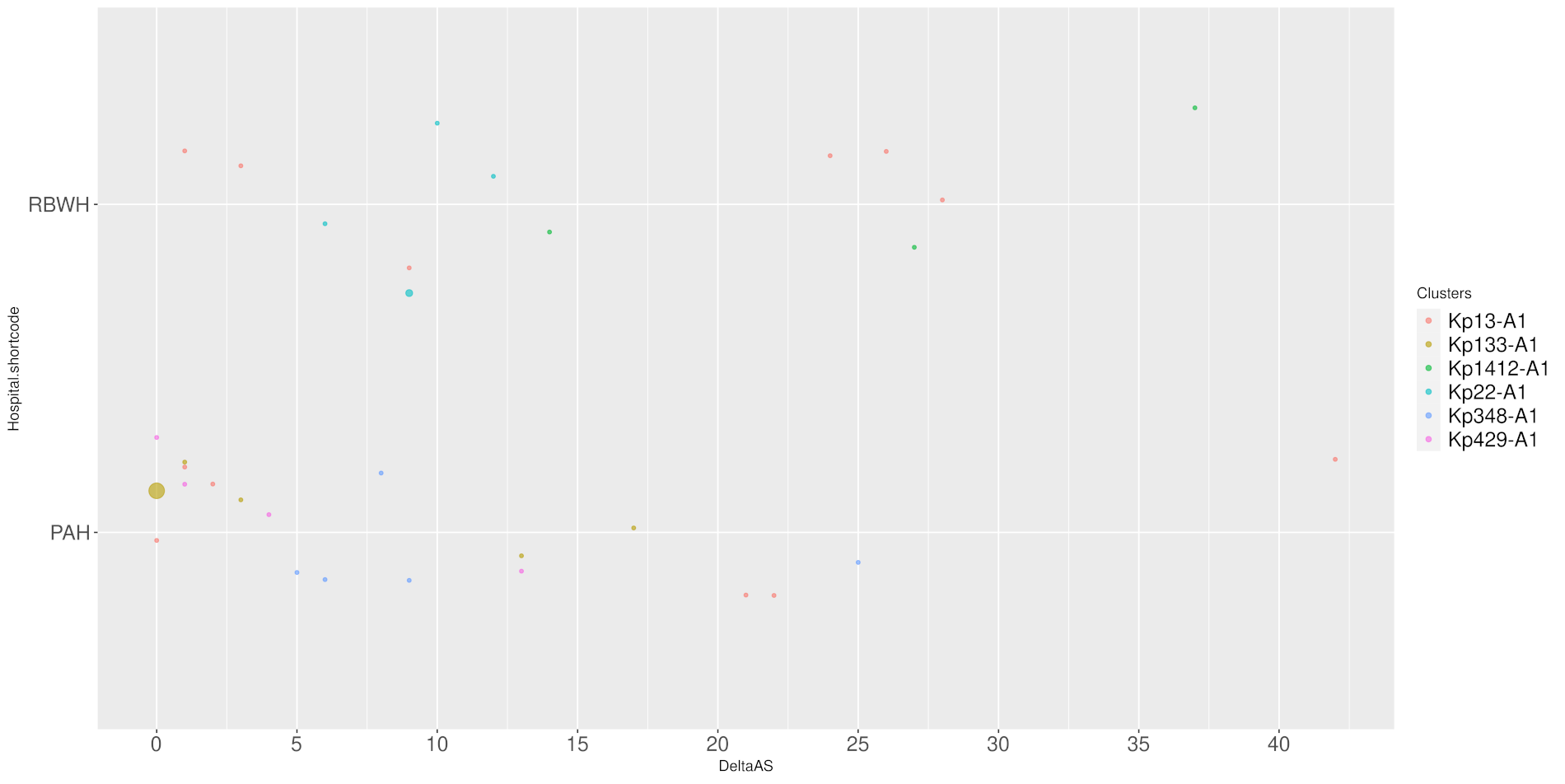

Figure S5: Patient sampling, days post admission - K. pneumoniae.

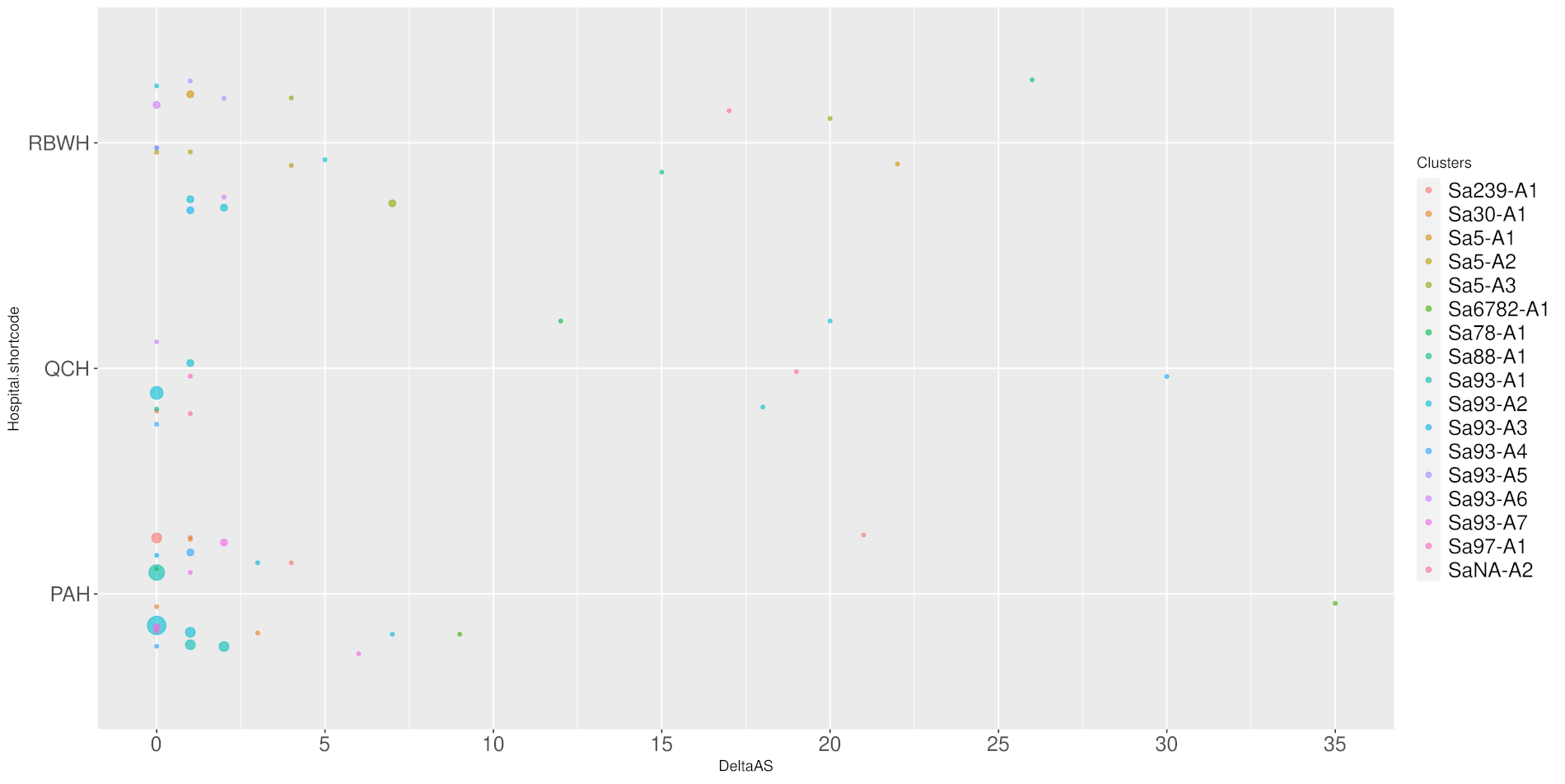

Figure S6: Patient sampling, days post admission - S. aureus.

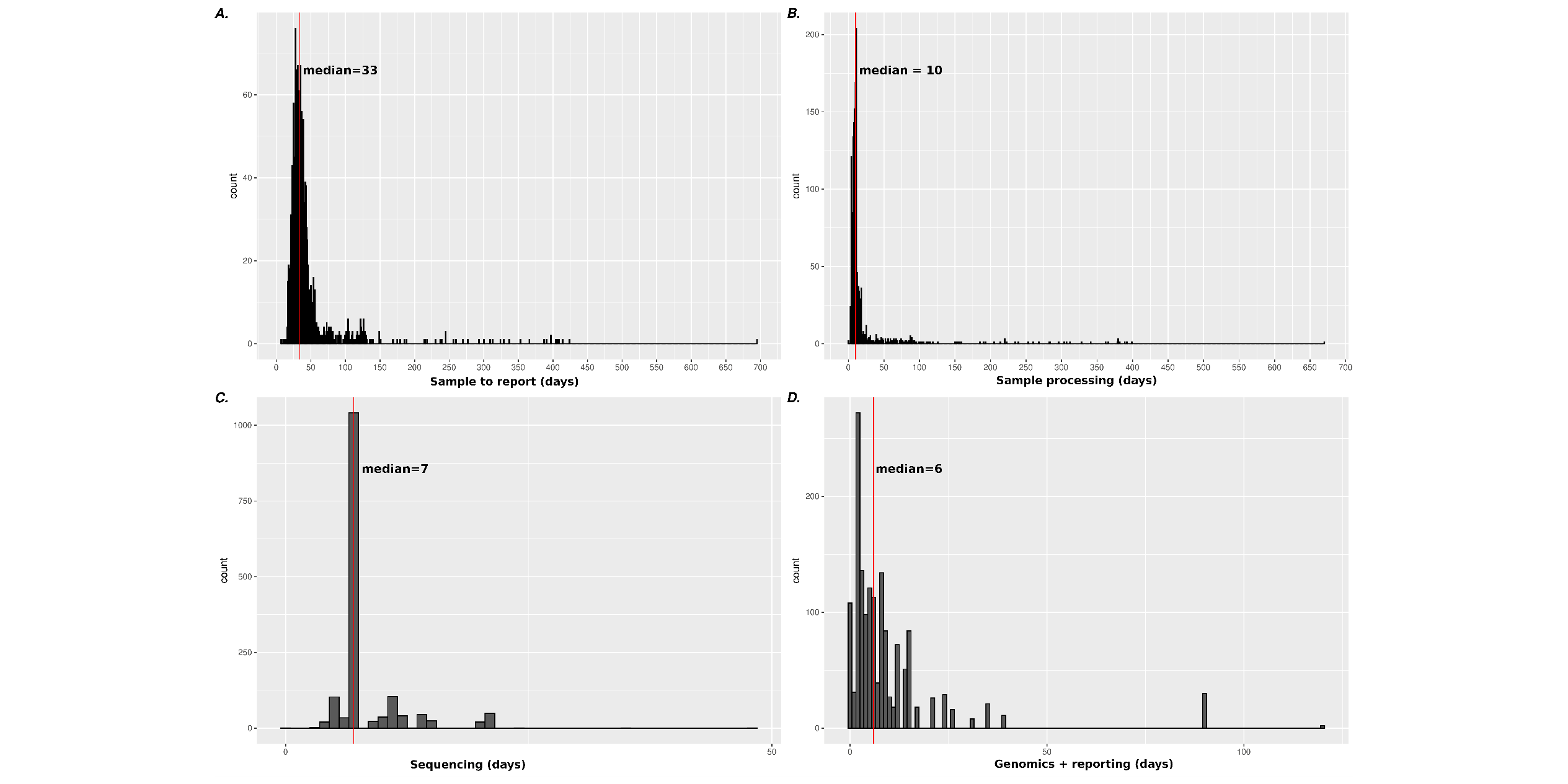

Figure S7: Turn-around times for (A) sample collection to reporting; (B) Sample collection to sequencing; (C) sample processing to sequencing; (D) sequencing to report generation; Red line = median duration (days)
