## Supplementary material - Surveillance report template for "Clinical implementation of routine whole-genome sequencing for hospital infection control of multi-drug resistant pathogens"

### Multi-Resistant Organism Genome Sequencing Report

Queensland Genomics Health Alliance

**Date:**

**<Report TITLE>**

**Executive Summary:**

**1. Isolates:**

**Table1: Sample information**

| Sample ID | URN | Lab No. | sample date | site | Species | MLST (ST) | Hospital | Admission date | Ward |
|---|---|---|---|---|---|---|---|---|---|

MLST = Multi-Locus Sequence Typing; NF =Not found;

**2. Cluster detection:**

*See cathai for clustering information (https://cathai.beatsonlab.com)*

**This reporting period:**

**QGHA survey period:**

**Public databases:** not tested.

***Clustered** isolates share **5 core genome SNPs per Mb**. Other hospitals include RBWH, PAH, QCH (LCCH). QGHA survey period began 01/11/18*

This report is not NATA accredited and is for **research use only**

### Multi-Resistant Organism Genome Sequencing Report

Queensland Genomics Health Alliance

#### Date:

#### 3. Resistance gene profiling:

**Table 2: Resistance gene profiles**

| Name | Species | MLST (ST) | Hospital | Gene[1] | Gene[2] | … | Gene[N] |
|---|---|---|---|---|---|---|---|

#### Comments:

#### Methods:

**Analysis:** <Name>
**Report:** <Name>

This report is not NATA accredited and is for **research use only**
