## Supplementary material - Patient report template for "Clinical implementation of routine whole-genome sequencing for hospital infection control of multi-drug resistant pathogens"

**GENOME SEQUENCING REPORT**
Research only – Not for diagnostic use

Queensland Genomics

### Patient info

| **Report requested by:** | <login ID> | **Date:** | <Date of request> |
|---|---|---|---|

| | | | |
|---|---|---|---|
| **Lab number** | <lab No> | **Hospital** | <Hospital> |
| **URN** | <URN> | **Ward** | <Ward> |
| **DOB** | <day/month/year> | | |

### Sample info

| **Sample no** | **Sample ID** | **Organism** | **Sample date** | **Isolation site** |
|---|---|---|---|---|
| Max 3/patient | <Sample ID> | <Species> | <Sample date> | <Site> |

*Other samples from same patient*

| | | |
|---|---|---|
| 1 | <Lab No> | 3 |
| 2 | NA if none | |

### Cluster Detection

**Summary:** Isolate <Lab No> clusters/ does not cluster with previously sequenced isolates

**Cluster ID:** <Cluster ID> Possible include cathai image here

**Caveat:** ***Clustered*** *isolates are assumed to share* ***≤ 5 core genome SNPs per Mb****.*

### Antibiotic Resistance Genes

| **Resistance Profile** | | | |
|---|---|---|---|
| **Gram-negatives** | **<Resistance profile – Gram-negatives>** | **Gram-positives** | **<Resistance profile – Gram-positives>** |

| **Identified Antibiotic/antimicrobial Resistance Genes** | |
|---|---|
| **Aminoglycosides** | |
| **Beta-lactamases** | |
| *Narrow* | TEM-1;OXA-1;etc;etc |
| *Broad* | |
| *ESBL* | |
| *Carbapenamase* | |
| *AmpC-Type* | |
| *Unknown* | |
| **Fluroquinolones** | |
| **etc** | |
| **etc** | |
